# Polygenic risk scores for psychiatric, inflammatory, and cardio-metabolic traits and diseases highlight possible genetic overlaps with suicide attempt and treatment-emergent suicidal ideation

**DOI:** 10.1101/2021.03.08.21253145

**Authors:** Giuseppe Fanelli, Marcus Sokolowski, Danuta Wasserman, European College of Neuropsychopharmacology (ECNP) Network on Suicide Research and Prevention, Siegfried Kasper, Joseph Zohar, Daniel Souery, Stuart Montgomery, Diego Albani, Gianluigi Forloni, Panagiotis Ferentinos, Dan Rujescu, Julien Mendlewicz, Diana De Ronchi, Alessandro Serretti, Chiara Fabbri

**Author notes:** Corresponding author*: Alessandro Serretti, MD, PhD, Department of Biomedical and Neuromotor Sciences University of Bologna, Viale Carlo Pepoli 5, 40123 Bologna, Italy.

## Abstract

Suicide is the second leading cause of death among young people. Genetics may contribute to suicidal phenotypes and their co-occurrence in other psychiatric and medical conditions. Our study aimed to investigate the association of polygenic risk scores (PRSs) for 22 psychiatric, inflammatory, and cardio-metabolic traits and diseases with suicide attempt (SA) or treatment-worsening/emergent suicidal ideation (TWESI).

PRSs were computed based on summary statistics of genome-wide association studies. Regression analyses were performed between PRSs and SA or TWESI in four clinical cohorts, including up to 3,834 individuals, and results were meta-analyzed across samples. Stratified genetic covariance analyses were performed to investigate the biology underlying cross-phenotype PRS associations. After Bonferroni correction, PRS for major depressive disorder (MDD) was positively associated with SA (p=1.7e-4). Nominal associations were shown between PRSs for coronary artery disease (CAD) (p=4.6e-3) or loneliness (p=0.009) and SA, PRSs for MDD or CAD and TWESI (p=0.033 and p=0.032, respectively). Genetic covariance between MDD and SA was shown in 35 gene sets related to drugs having anti-suicidal effects.

A higher genetic liability for MDD may underlie a higher risk of SA. Further, but milder, possible modulatory factors are genetic risk for loneliness and CAD.

## 1. Introduction

It has been estimated that one person every 40 seconds dies from suicide worldwide, resulting in ∼800,000 deaths per year. Suicide is the second most prevalent cause of death among young people, with European countries having the highest death rates [WHO 2019]. To address this dramatic epidemic, the World Health Organization (WHO) has included the reduction of suicide mortality rate among the explicit targets for 2030, to be reached by implementing prevention strategies and promoting mental health and healthy lifestyles.

Suicidal phenotypes are ten times more prevalent among patients suffering from psychiatric disorders than in the general population, with a prevalence of mental illness of ∼90% among people who commit/attempt suicide. Among these, largely common are mood disorders and schizophrenia as well as substance use disorders [Arsenault-Lapierre and others 2004]. About 10% of depressed patients may also experience the emergence or worsening of suicidal ideation in the early stages of antidepressant therapy [Cristancho and others 2017]. Indeed, despite lengthy controversy in the scientific community, the US Food and Drug Administration maintains a black box warning regarding the increased risk of suicidal thoughts in people <25 years of age being treated with antidepressants, and still recommends close monitoring in all other age groups regarding this possible risk [Friedman 2014]. Less attention has been paid to the association of physical comorbidities with suicidal phenotypes. However, it is known that inflammatory and cardio-metabolic diseases are associated with increased suicide risk, regardless of the co-occurrence of any mental disorders, and that suicidal events are often preceded by access to primary care for physical complaints [Luoma and others 2002; Stickley and others 2020].

The biopsychosocial model represents the most widely accepted theory of the onset of suicidal behaviors. It considers the suicidal event as the result of the interplay between distal (or predisposing) factors, consisting of genetic/epigenetic influences and early life adversities leading to lasting alterations in gene expression, and proximal (or precipitating) factors [Turecki and others 2019], which include recent socio-economic changes and stressful life events. Among proximal factors, social isolation and perceived loneliness have been shown to be important [Calati and others 2019]. Personality traits represent key mediators between distal and proximal factors. In this regard, impulsive-aggressive traits are among the most frequently associated with suicide and are often found in externalizing disorders like attention-deficit/hyperactivity disorder (ADHD) [Nock and others 2009]. In addition, anxiety traits/disorders and impaired self-control have been closely related to the transition from suicidal ideation to suicide attempts (SA) [Nock and others 2009]. However, the etiopathogenesis of suicide is far from being fully understood. Genetic factors have been shown to play a role in suicidal behaviours, with heritability from 30% to 55% in twin and adoption studies, whereas a more precise estimate could be 17-36% once the heritability of comorbid psychiatric conditions is accounted for [Fanelli and Serretti 2019; Fu and others 2002; Voracek and Loibl 2007]. However, SNP-based heritability (h^2^_SNP_) of SA was only ∼4% in genome-wide association studies (GWAS), but more insights are likely to come as sample sizes increase [Mullins and others 2019; Ruderfer and others 2020].

A promising research opportunity is represented by polygenic risk scores (PRSs), which summarize the additive genetic risk conferred by multiple common variants across the genome [Choi and O’Reilly 2019]. As well as being promising tools for stratifying individuals at higher risk, PRSs also allow the investigation of possible shared genetics between complex traits [Fanelli and others 2020], therefore they can provide insights on the genetic overlap between suicidal phenotypes and other psychiatric or physical conditions. Further information may derive from stratifying the genome by functional annotations [Lu and others 2017], which could lead to a deeper understanding and the formulation of new hypotheses about the biological mechanisms underlying suicidal phenotypes.

Hence, this study aimed to: (1) test the association between the PRSs for 22 psychiatric, inflammatory, and cardio-metabolic traits/diseases with SA in major psychiatric disorders or treatment-worsening/emergent suicidal ideation in depression (TWESI); (2) investigate the common molecular pathways underlying cross-phenotype genetic overlaps by exploring pairwise genetic covariance stratified at the level of candidate gene sets, which were selected as those involved in the response/pharmacodynamics of psychotropic drugs with known anti-suicidal effects (i.e., clozapine, ketamine, and lithium).

## 2. Methods

### 2.1. Target samples

#### Clinical Antipsychotic Trials of Intervention Effectiveness (CATIE)

The CATIE study is a randomized clinical trial sponsored by the National Institute of Mental Health (NIMH) intended to assess the efficacy of one first-generation and four second-generation antipsychotic medications in patients with schizophrenia. Eligible patients were initially randomized under double-blind conditions to perphenazine, olanzapine, quetiapine, risperidone, or ziprasidone and received medication for up to 18 months or before discontinuation of treatment for any cause. Additional information can be found elsewhere [Stroup and others 2003].

#### European Group for the Study of Resistant Depression (GSRD)

The GSRD is a multicentre study designed to investigate antidepressant treatment response/resistance in patients with major depressive disorder (MDD). Patients were excluded if they were diagnosed with any other primary psychiatric disorder or substance use disorder in the previous six months. Treatment with antidepressants was conducted in a naturalistic manner following the best clinical practice (each antidepressant was taken at an appropriate dose for ≥4 weeks during each depressive episode). Additional details can be found elsewhere [Dold and others 2018].

#### Sequenced Treatment Alternatives to Relieve Depression (STAR*D)

The STAR*D study was conducted to evaluate the efficacy and tolerability of different antidepressant therapies across four sequential treatment stages in patients with MDD. Patients with non-psychotic MDD (DSM-IV criteria) were recruited from primary care or outpatient psychiatric services. The study design and population are described in depth elsewhere [Howland 2008].

### Systematic Treatment Enhancement Program for Bipolar Disorder (STEP-BD)

STEP-BD was a prospective study aimed at improving bipolar disorder treatment and management and evaluating the longitudinal outcome of the disorder. A hybrid design was used by STEP-BD to gather longitudinal data as patients move between naturalistic studies and randomized controlled trials. Patients aged 15 yo with type I or II bipolar disorder (BIP), cyclothymia, BIP NOS, cyclothymia or schizoaffective disorder were enrolled. The research design and study population are described in depth elsewhere [Sachs and others 2003].

### 2.2. Target phenotypes

Two binary target phenotypes were considered: suicide attempt (SA), which was extracted from all the four target samples, and treatment-worsening/emergent suicidal ideation (TWESI), which was available in the two MDD samples (GSRD and STAR*D).

SA was defined as an intentional, self-injurious behavior with some potential for serious harm and/or lethality [Dennehy and others 2011; Marangell and others 2008]. In more detail, in CATIE SA was assessed based on the Calgary Depression Rating Scale (CDRS) item 8 (“Suicide attempt”), the Family/Caregiver Interview (FI) item 78 (“Attempted suicide during the past 6 months”) at the baseline and follow-up. In GSRD, SA was defined according to the 21-item HDRS (Hamilton Depression Rating Scale) item 3 (“Suicide”) or MINI (Mini-International Neuropsychiatric Interview) items C5-C6a (“In the past month did you attempt suicide?”, “In your lifetime did you ever make a suicide attempt?”). In STAR*D, SA was assessed through the HDRS item 18 (“Suicide”), the screening CRF (Case Report Form) item 21 (“Attempted suicide”), SAE (Serious Adverse Event) form item 19 (“Failed suicide attempt”), IVR (Interactive Voice Response) Alert form (“Primary reason for alert: suicide attempt”). In STEP-BD, SA was derived from the SQ (Suicide Questionnaire) item 3 (“Tried to hurt yourself or attempt suicide”), SAE (Serious Adverse Experience) report (“Was there a suicide attempt?”), ADE (Affective Disorders Evaluation) form (“History of suicide attempts”), Care Utilization (CU) form (“Suicide attempts, last 3 months”). In line with the previous literature [Laje and others 2007; Perlis and others 2007], in GSRD the TWESI phenotype was derived as a Δ≥1 change from baseline in MADRS (Montgomery Asberg Depression Rating Scale) item 10 score and in STAR*D as a Δ≥1 change from baseline in the QIDS-SR16 (16-item Quick Inventory of Depressive Symptomatology – Self-Report) item 12.

### 2.3. Genotyping and quality control (QC) of the target samples

Individuals in the CATIE study were genotyped by Perlegen Sciences (Mountain View, CA, USA) using the Affymetrix GeneChip Human Mapping 500K Array Set (Santa Clara, CA, USA), comprising Nsp and Sty chips, and a custom 164K chip built by Perlegen to obtain a wider genome coverage. Genotyping of the GSRD participants was conducted using the Illumina Infinium PsychArray-24 BeadChip (San Diego, CA, USA). Individual-level genotypes in STAR*D were obtained using the Affymetrix GeneChip Human Mapping 500K Array Set or Affymetrix Genome-Wide Human SNP Array 5.0. For STEP-BD, genotyping was carried out by the Genetic Analysis Platform at the Broad Institute (Cambridge, MA, USA) using the Affymetrix GeneChip Human Mapping 500K Array Set. Pre-imputation QC was carried out in all the target samples by first removing monomorphic variants and the single-nucleotide polymorphisms (SNPs) with a genotype missing rate ≥5%. Individuals who were non-European or had sex mismatches, a genotyping rate <97%, abnormal heterozygosity, high relatedness (identity by descent (IBD)>0.1875) (Anderson et al. 2012) were excluded. Population principal components were calculated using a linkage disequilibrium-pruned set of variants (R^2^<0.2) and individuals falling outside ±5 standard deviations from the mean of the top 20 population principal components were excluded. Genotype imputation was performed using Minimac3 and the Reference Consortium (HRC) r1.1 2016 reference panel. Variants with minor allele frequency (MAF) <0.01, low imputation accuracy (r^2^ (estimated squared correlation between imputed genotypes and true genotypes) <0.30) [Li and others 2010], and genotype probability <0.9 were removed.

### 2.4. Statistical analysis

#### 2.4.1. Polygenic risk scores (PRSs)

For each individual in the target samples, PRSs for up to 22 psychiatric, inflammatory, and cardio-metabolic traits/diseases were computed as the sum of the number of effect alleles at each SNP position, weighted for their effect size derived from the largest GWAS meta-analyses available at the time of conducting our analyses. A complete list and further information on base samples is reported in Table 1.

**Table 1.** Base samples used for the computation of the genome-wide polygenic risk scores. Abbreviations: N, total sample size; Neff, effective sample size [Neff=4/(1 /(N cases)+1/(N controls)].

| **Base trait/disorder** | **Reference** | **Year** | **PMID** | **N cases** | **N controls** | **N** | **Neff** |
| --- | --- | --- | --- | --- | --- | --- | --- |
| *ADHD (attention-deficit/hyperactivity disorder)* | Demontis et al. | 2019 | 30478444 | 19,099 | 34,194 | **53,293** | 49,017 |
| *Aggression (childhood and early adolescence aggressive behavior)* | Pappa et al. | 2018 | 26087016 |  |  | **18,988** |  |
| *Agreeableness* | De Moor et al. | 2010 | 21173776 |  |  | **17,375** |  |
| *Alcohol dependence* | Walters et al. | 2018 | 30482948 | 11,569 | 34,999 | **46,568** | 34,780 |
| *Alcohol Intake (drinks per week)* | Liu et al. | 2019 | 30643251 |  |  | **537,349** |  |
| *Anxiety (lifetime anxiety disorder)* | Purves et al. | 2019 | 31748690 | 25,453 | 58,113 | **114,019** | 70,802 |
| *BIP (bipolar disorder)* | Stahl et al. | 2018 | 29906448 | 20,352 | 31,358 | **51,710** | 49,367 |
| *CAD (coronary artery disease+total cholesterol)* | Siewert et al. | 2018 | 30525989 |  |  | **735,838** |  |
| *Cannabis (lifetime Cannabis Use)* | Pasman et al. | 2018 | 30150663 | 43,380 | 118,702 | **162,082** | 127,079 |
| *Conscientiousness* | De Moor et al. | 2010 | 21173776 |  |  | **17,375** |  |
| *CRP (C-reactive protein) - chronic low-grade inflammation* | Ligthart et al. | 2018 | 30388399 |  |  | **204,402** |  |
| *Extraversion* | Van den Berg et al. | 2016 | 26362575 |  |  | **63,030** |  |
| *Insomnia* | Jansen et al. | 2019 | 30804565 | 109,402 | 277,131 | **386,533** | 313,750 |
| *Loneliness* | Day et al. | 2018 | 29970889 | 80,134 | 364,890 | **445,024** | 262,818 |
| *MDD (major depressive disorder)* | Wray et al./Howard et al. | 2018/2019 | 29700475/30718901 | 170,756 | 329,443 | **500,199** | 449,856 |
| *MetS (metabolic syndrome)* | Lind | 2019 | 31589552 | 59,677 | 231,430 | **291,107** | 189,773 |
| *Neuroticism* | Baselmans et al. | 2019 | 30643256 |  |  | **523,783** |  |
| *Openness to experience* | De Moor et al. | 2010 | 21173776 |  |  | **17,375** |  |
| *PGC2-cross-disorder phenotype* | PGC-Cross Disorder Group 2 | 2019 | 31835028 | 232,964 | 494,162 | **727,126** | 633,299 |
| *SCZ (schizophrenia)* | Ripke et al. | 2014 | 25056061 | 36,989 | 113,075 | **150,064** | 111,487 |
| *SA (suicide attempt)* | Ruderfer et al. | 2019 | 30610202 | 2,433 | 334,766 | **157,366** | 9,662 |
| *T2DM (type 2 diabetes mellitus)* | Mahajan et al. | 2018 | 30297969 | 74,124 | 824,006 | **898,130** | 272,026 |

In each target sample, PRSs for each base trait/disease were calculated using PRSice-2.2.13 at 8 *a priori* GWAS P-thresholds (P_T_) (1e-4, 0.001, 0.05, 0.1, 0.2, 0.3, 0.4, 0.5) [Choi and O’Reilly 2019]. SNPs in high LD were clumped considering a 250kb window and r^2^ threshold of 0.1. Logistic regression analyses between PRSs for each trait/disease and the case-control status (i.e., SA *vs* non-SA, TWESI *vs* non-TWESI) were performed in each target sample, adjusting for population stratification and recruitment centers. The proportion of variance in SA or TWESI explained by PRSs in each sample was estimated by Nagelkerke’s pseudo-R^2^ as the difference between the R^2^ of the full model, incorporating the PRS and covariates, and the R^2^ of the null model, including only the covariates. Finally, results of PRSs analyses at each P_T_ across samples were meta-analyzed for both the target phenotypes using a fixed-effect inverse-variance weighted model with the metafor R-package (https://cran.r-project.org/web/packages/metafor), as done by previous authors [Garcia-Gonzalez and others 2017; Zheutlin and others 2019]. Between-study heterogeneity was assessed using the Cochran’s Q test, computed as the weighted sum of the squared differences between the individual and pooled study effects, and Higgin’s and Thompson’s I^2^, describing the percentage of variation across studies being due to heterogeneity rather than chance [Higgins and others 2003]. Bonferroni correction was applied to account for the multiple base traits/diseases and the eight P_T_ considered for the PRSs analyses (α=0.05/(22*8)=2.8e-4).

The statistical power of PRSs was determined via the AVENGEME R-package [Palla and Dudbridge 2015]. We assumed a covariance between the genetic effects in the base and target samples of 25 or 50% [Garcia-Gonzalez and others 2017], while sample and lifetime population prevalences, h^2^ _SNP_ of base and target phenotypes were derived from the previous literature (Table S1). The PRSs of all the 22 base traits/diseases showed adequate predicting power (>80%) for SA and TWESI.

#### 2.4.2. Genetic covariance analysis stratified by candidate gene sets

To better characterize the biology underlying the genetic overlap of the considered psychiatric and non-psychiatric traits with suicidal phenotypes, we performed pairwise genetic covariance analyses stratified by gene sets using GeNetic cOVariance Analyzer (GNOVA), a tool that provides genetic covariance estimates that are robust to sample overlap [Lu and others 2017]. As starting point, we considered the biological mechanisms involved in the effects of drugs with well-known anti-suicidal effects (i.e., clozapine, ketamine, lithium), as these have strong evidence of being implicated in suicidal phenotypes. We reviewed the previous literature looking for genes and molecular pathways implicated in the effect of these drugs (Table S2). The mechanisms and genes identified by the literature review were used as search queries in Molecular Signatures Database (MSigDB) v7.2 (https://www.gsea-msigdb.org/gsea/msigdb/index.jsp), a public collection of functionally annotated gene sets, filtering for hallmark, curated (e.g., BioCarta, Broad Institute, Kyoto Encyclopedia of Genes and Genomes (KEGG)), and Gene Ontology (GO) gene sets in Homo Sapiens; two-hundred-seventeen relevant gene sets were identified.

Each gene-set was annotated to SNP positions on the 1k Genomes Project Phase 3 (1kGP3) reference panel by using the LDSC (Linkage Disequilibrium Score regression) *make_annot*.*py* function [Bulik-Sullivan and others 2015]. For the PRSs associated with suicidal phenotypes, the corresponding GWAS summary statistics were quality checked using the LDSC munging function, removing SNPs with a MAF ≤0.01 (when MAF was available) and those not matching HapMap3 SNPs, as these are generally well-imputed; SNPs having missing or out-of-bounds values (ranges for INFO 0-1.5; MAF: 0-1; p-values: 0-1), and all indels, structural and strand-ambiguous variants were also pruned. Munged GWAS summary statistics of the phenotypic pairs under investigation were used as input datasets for the genetic covariance analyses. Bonferroni correction was applied (α=0.05/217 gene sets=2.3e-4).

In order to prioritize the results obtained in GNOVA, we performed partitioned heritability analyses in LDSC by assessing the h^2^_SNP_ of SA at the level of the individual tested gene sets [Finucane and others 2015]. The same munged summary statistics used in the GNOVA analyses as well as LD scores partitioned for each gene-set were used as input in LDSC for these analyses.

## 3. Results

After QC of target samples, a total of 3,834 patients were included in the PRS meta-analyses for SA, of which 688 were cases. Of these 3,844 subjects, 478 were diagnosed with schizophrenia, 2,601 with MDD, and 755 with disorders of the bipolar spectrum. The PRS meta-analyses on TWESI included a total of 2,574 patients with MDD from the STAR*D and GSRD samples, of whom 214 subjects were cases. Further details on the number of cases/controls in the four samples are shown in Table 2.

**Table 2.** Number of individuals showing either SA or TWESI in the four clinical cohorts included in our analyses. Abbreviations: SA, suicide attempts; TWESI, treatment-worsening-emergent suicidal ideation; CATIE, Clinical Antipsychotic Trials of Intervention Effectiveness; GSRD, European Group for the Study of Resistant Depression; STAR*D, Sequenced Treatment Alternatives to Relieve Depression, STEP-BD, Systematic Treatment Enhancement Program for Bipolar Disorder.

|  | **SA** | **non-SA** | **SA + non-SA** |
| --- | --- | --- | --- |
| **CATIE** | 23 | 455 | 478 |
| **GSRD** | 112 | 1,037 | 1,149 |
| **STAR\*D** | 211 | 1,241 | 1,452 |
| **STEP-BD** | 342 | 413 | 755 |
| **total** | **688** | **3146** | **3,834** |

|  | **TWESI** | **non-TWESI** | **TWESI + non-TWESI** |
| --- | --- | --- | --- |
| **GSRD** | 129 | 1,020 | 1,149 |
| **STAR\*D** | 85 | 1,340 | 1,425 |
| **total** | **214** | **2,360** | **2,574** |

### Association between PRSs for MDD and SA across samples

After Bonferroni correction, our meta-analyses highlighted a positive association between MDD-PRS and SA at P_T_ =0.05 (pooled β=0.212 (95% CI 0.101-0.322; p=1.7e-4; R^2^=0.4%-1.6%). The direction of the association effect between MDD-PRS and SA was concordant across the four target samples, and no heterogeneity was detected (χ^2^_QE_ =2.49, QE p=0.476; I^2^=0%; Fig. S1 shows a forest plot of the MDD-PRS meta-analysis on SA). The largest effect was found in the CATIE sample albeit with a large standard error, while the most significant effect was found in the STAR*D sample, where all the tested P-thresholds showed nominally significant associations (p=0.04-9e-4, R^2^=0.5%-1.3%). Across the four included cohorts, patients in the 5th MDD-PRS quintile showed a higher risk of SA than individuals in the lowest quintile after the meta-analysis (OR=2, 95% CI 1.42-2.80, p=7.27e-5). PRS meta-analyses also showed nominal associations between SA and the PRSs for coronary artery disease (CAD) (p=4.6e-3), loneliness (p=0.009), and SA (p=0.034) (Fig. 1 and Table S3). No PRS was associated with TWESI after multiple-testing correction, although MDD- and CAD-PRS showed nominal associations with TWESI in the meta-analysis (p=0.033 and p=0.032, respectively) (Fig. 2 and Table S4).

**Figure 1.**
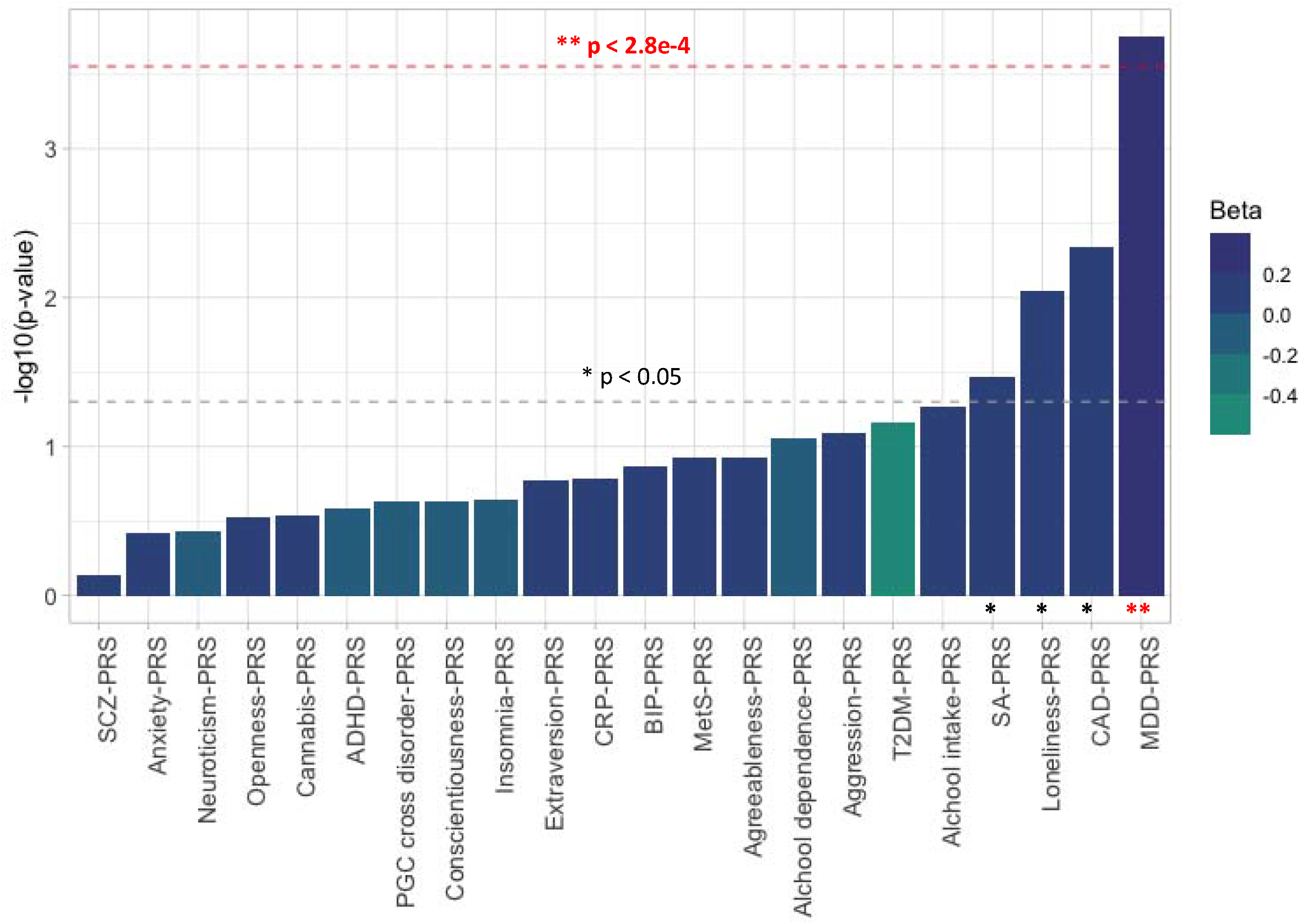
Bar plot showing the associations between the PRSs for multi psychiatric, inflammatory and cardio-metabolic diseases/traits and **suicide attempt** (**SA**) in an overall case-control sample (after the meta-analysis) of 3,834 individuals suffering from major depressive disorder, bipolar disorder spectrum or schizophrenia. Best-fitting PRSs are depicted in increasing order of significance (-log10 p-values) of association with SA. The red dashed line corresponds to the Bonferroni corrected threshold of statistical significance (α=0.05/(22*8)=2.8e-4). The gray dashed line indicates the nominal threshold of significance (p=0.05). Abbreviations: ADHD, attention-deficit/hyperactivity disorder; BIP, bipolar disorder; CAD, coronary artery disease; CRP, C-reactive protein; MDD, major depressive disorder; MetS, metabolic syndrome; T2DM, type 2 diabetes mellitus; SCZ, schizophrenia.

**Figure 2.**
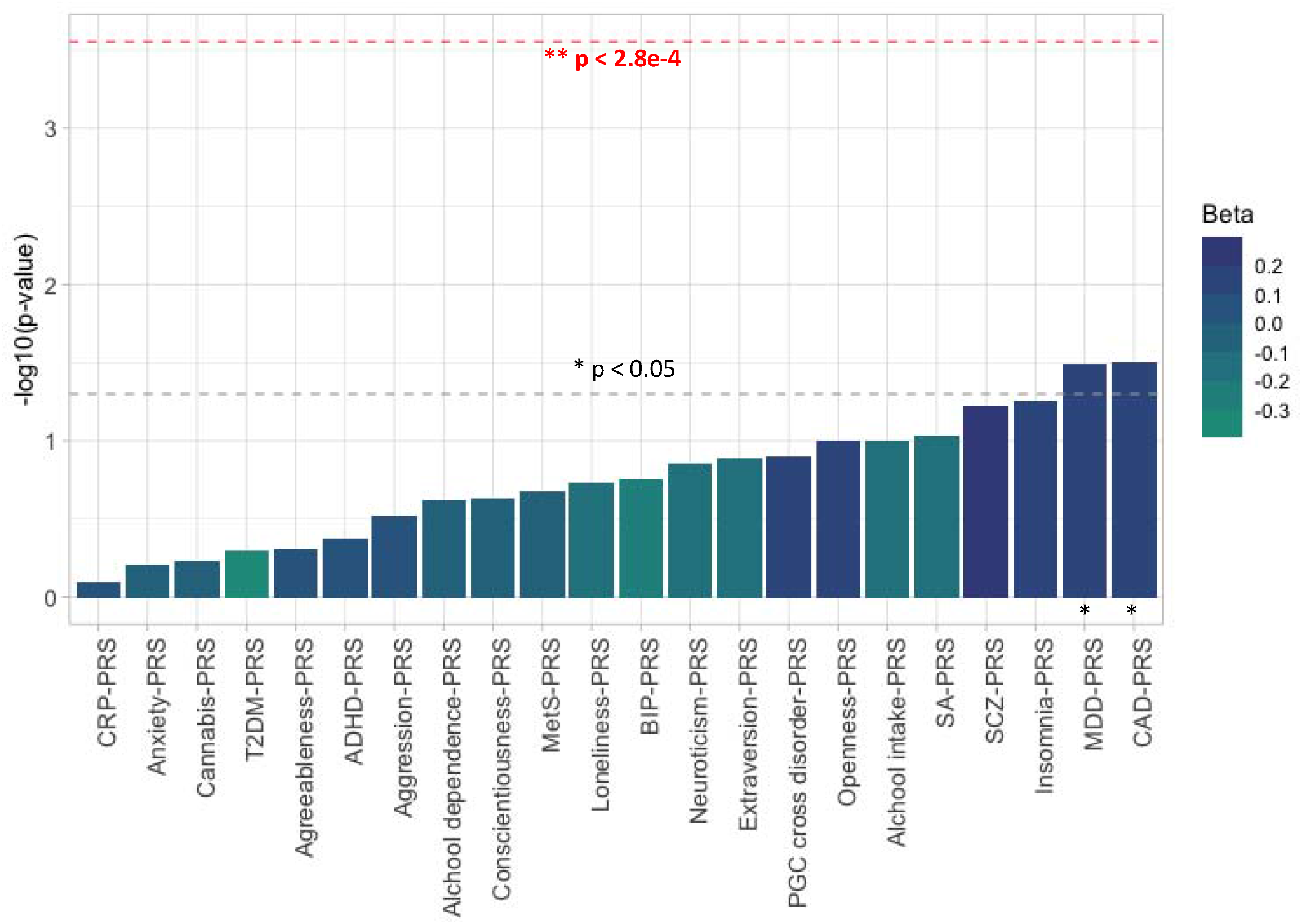
Bar plot showing the associations between the PRSs for multi psychiatric, inflammatory and cardio-metabolic traits/diseases and the **treatment-emergent/worsening suicidal ideation** (**TWESI**) phenotype in an overall case-control sample (after the meta-analysis) of 2,574 individuals suffering from MDD. Best-fitting PRSs are depicted in increasing order of significance (-log10 p-values) of association with TWESI. The red dashed line corresponds to the Bonferroni corrected threshold of statistical significance (α=0.05/(22*8)=2.8e-4). The gray dashed line indicates the nominal threshold of significance (p=0.05). Abbreviations: ADHD, attention-deficit/hyperactivity disorder; BIP, bipolar disorder; CAD, coronary artery disease; CRP, C-reactive protein; MDD, major depressive disorder; MetS, metabolic syndrome; T2DM, type 2 diabetes mellitus; SCZ, schizophrenia.

### Genetic covariance between MDD and SA stratified by gene sets

After correcting for multiple testing, functionally stratified GNOVA analyses revealed significant genetic covariance between MDD and SA at the level of 35 gene sets (p<2.24e-4; Table S5). We prioritized these gene sets by checking their partitioned SA h^2^_SNP_ and we found a significant h^2^_SNP_ for Reactome tumor necrosis factor receptor-1 (TNFR1)-induced proapoptotic signaling and GO α-adrenergic receptor activity (p<2.24e-4); however, only the h^2^ _SNP_ estimate for GO α-adrenergic receptor activity had a value >0 (h^2^_SNP_=8e-4; p=6.33e-5) (Table S5).

## 4. Discussion

In this study, we investigated whether PRSs for multiple psychiatric, inflammatory, and/or cardio-metabolic diseases and traits were associated with SA or TWESI in a pooled sample of up to 3,834 patients with mood disorders or schizophrenia. Our results suggested that a higher genetic liability for MDD underlies an increased risk of SA among patients with psychiatric disorders. PRSs for SA and loneliness were found to be nominally associated with SA across samples, as well as MDD-PRS with TWESI in patients with MDD. This was the first study to investigate whether the PRSs for inflammatory and cardio-metabolic traits/diseases are associated with SA or TWESI. According to our findings, the impact of the genetic risk for inflammatory and cardio-metabolic diseases/traits was limited for the examined suicidal phenotypes, although trends of association between CAD-PRS and SA or TWESI were shown.

Our findings are consistent with previous evidence of shared genetics between MDD and SA, as other authors have previously suggested through PRSs and bivariate genetic correlation analyses [Mullins and others 2019; Ruderfer and others 2020]. An association between MDD-PRS and SA had been demonstrated in patients with MDD, bipolar disorder and schizophrenia (p=2e-4 - 6e-4; R^2^=0.24-0.40%) by using a base dataset for MDD approximately three times smaller (n=∼171,000) and independent target samples than those used in our analyses [Mullins and others 2019]. Of note, our results also showed a more significant effect of MDD-PRS on SA in patients with MDD than in those affected by schizophrenia or mood swings, although the association signal remained concordant across the four considered target samples and significant in the meta-analysis (with a variance in the target phenotype explained by MDD-PRS of up to 1.6%). The genetic overlap with MDD may not be confined to the SA phenotype only, but also to TWESI, in line with previous studies [Mullins and others 2014], though the association with TWESI did not survive Bonferroni correction.

We further investigated the genetic overlap between MDD and SA using GNOVA and 217 gene sets relevant for the biological mechanisms involved in suicide; the results showed significant genetic covariance at the level of 35 gene sets. Among these, the majority (i.e., 27/35) showed a concordant direction of effect between MDD and SA, as expected from the PRS analyses and previous epidemiological evidence. For the gene sets showing significant genetic covariance, the partitioned h^2^ _SNP_ of SA supported a positive and significant genetic component only for the α-adrenergic receptor activity gene-set (GO:0004936). α_2_-adrenoreceptors are mostly auto-/hetero-receptors located at the presynaptic level and negatively regulate the release of neurotransmitters such as norepinephrine and serotonin [Aoki and others 1998]. Replicated evidence demonstrated an up-regulation and increased activity of α_2_-adrenoreceptors in the hippocampus and prefrontal cortex of MDD patients who died by suicide; in these individuals, α_2_-adrenoreceptors seem not affected by the expected down-regulatory effects of antidepressants [Rivero and others 2014]. Interestingly, noradrenergic neurotransmission also plays a role in aggression, which is considered an endophenotype of suicide, and higher plasma levels of norepinephrine have been associated with decreased aggressive and suicidal behaviours [Yanowitch and Coccaro 2011]. The role of α_2_-adrenoreceptors in suicide is also supported by the observation that clozapine has the greatest differential affinity towards α-adrenoreceptors *vs* D_2_-receptors (the most potent affinity with respect to binding to the dopamine D_2_-receptor) among atypical antipsychotics [Stahl and Stahl 2013].

The lack of a positive and significant h^2^_SNP_ for SA in the other gene sets showing significant genetic covariance between SA and MDD may reflect a truly non-significant genetic component for SA in those pathways, but also be the consequence of sampling variation in small samples when the true heritability is near zero; in this regard, limited statistical power was suggested by previous GWAS of SA, since the large discrepancy between h^2^ _SNP_ (4%) and heritability found by twin studies (30-50%).

Among the nominally significant findings, we report that loneliness-PRS was associated with SA in patients with MDD. This finding appears relevant as loneliness is one of the core symptoms of depression, as well as being the depressive symptom that explains the greatest variance in suicidal ideation [Gijzen and others 2021]. Consistently, a recent meta-analysis supported a longitudinal association between loneliness, increased SA and depressive symptoms [Solmi and others 2020]. Therefore, targeting loneliness should be considered as a key goal in the treatment of MDD. We also reported a nominal association between CAD-PRS and both SA and TWESI, in line with the large amount of clinical evidence pointing to an increased risk of suicide in people with cardiovascular disease and *vice versa* [Artero and others 2006; Zhong and others 2020]. Although MDD has also been shown to be a risk factor for CAD, the association between CAD and suicide may be independent of depression risk, as suggested by previous studies [Artero and others 2006; Moazzami and others 2018]. The lack of association we found between C-reactive protein (CRP)-PRS and suicidal phenotypes might appear conflicting with the association shown between CAD-PRS and SA/TWESI, as epidemiological studies have indicated that increased peripheral CRP levels are present in patients with CAD [Strang and Schunkert 2014]. However, genomic studies have ruled out a role of CRP genetics in determining CAD, giving support to our observation [Strang and Schunkert 2014].

Our study comes with some strengths and limitations. The major strengths are the use of a panel of 22 traits/diseases, including inflammatory and cardio-metabolic phenotypes, for the construction of PRSs, as well as the use of a meta-analytic approach to summarize the results across samples and a strict Bonferroni correction that minimized type-1 errors. PRSs at different P_T_ are indeed correlated. We further investigated the biology underlying the association between MDD-PRS and SA through a genetic covariance analysis stratified by relevant gene sets. However, some limitations did not allow clear conclusions to be drawn. Among them, the small effective sample size (N_eff_, with a much lower number of cases than controls) and h^2^ _SNP_ explained in the SA GWAS used as reference for our GNOVA analyses [Ruderfer and others 2020], which made us cautious in interpreting the GNOVA results. Hence, these findings are currently to be considered as exploratory. Another limitation is that PRSs only consider additive SNP effects, while being unable to account for the effect of rare variants, epistatic or epigenetic mechanisms, which might also play a role in suicide phenotypes. Finally, it should be considered that the target samples were not primarily recruited to assess suicidal phenotypes.

In conclusion, our study highlighted that a higher genetic liability for MDD increases the risk of SA among patients with mood disorders and schizophrenia, pointing towards possible shared etiopathogenetic mechanisms between MDD and SA. In this regard, we suggested a convergent genetic signal at the level of the α-adrenoreceptor signalling pathway, in line with previous evidence linking this pathway with both MDD and suicide. Overall, our findings suggested limited or no genetic overlap between inflammatory and cardio-metabolic traits and suicidal phenotypes. Despite only nominally significant, the association between loneliness PRS and SA is consistent with previous strong evidence supporting the relevance of this trait on suicide risk. Therefore, the early and proper treatment of MDD, with a particular focus on feelings of loneliness, should be considered as pivotal to reduce suicide rates. These results may be useful for implementing algorithms that incorporate both clinical and genetic risk factors to identify individuals at higher risk for SA and thus eligible for targeted preventive campaigns.

### Statement of Ethics

Data were obtained for analysis from the National Institute of Mental Health (NIMH), Bethesda, Maryland, US (Request ID 5ce26a95712d8). The STAR-D, STEP-BD, and CATIE trials were conducted according to the Principles of Helsinki Declaration. The study protocol was reviewed and approved by ethical committees at local recruitment sites. All subjects selected by clinicians were included in the screening phase after obtaining their written informed consent. This research group certifies that data collected for the STAR-D, STEP-BD, and CATIE trials were exclusively used for scientific investigation. Before obtaining access to data, the objectives of our investigation were clearly described in the request form.

## Data Availability

-

## Acknowledgments

The European College of Neuropsychopharmacology (ECNP) Network on Suicide Research and Prevention commissioned this manuscript and contributed by acting as reference group and providing comments and critical review to the manuscript. The Chair of the Network is Professor Judith Balazs, Head of the Department of Developmental and Clinical Child Psychology at the Institute Psychology Eötvös Loránd University, and Director of Education and Research at the Vadaskert Child and Adolescent Psychiatric Hospital and Outpatient Clinic, Budapest, Hungary. The co-Chair is Professor Marcus Sokolowski, National Centre for Suicide Research and Prevention of Mental Ill-Health (NASP), Karolinska Institute, Stockholm, Sweden. Members of the Network are: Professor Alan Apter, Feinberg Child Study Centre, Schneider Children’s Medical Centre, Tel Aviv University, Tel Aviv, Israel; Professor Enrique Baca Garcia, Department of Psychiatry, Fundacion Jimenez Diaz University Hospital, Autonomous University of Madrid, Madrid, Spain; Professor Julio Bobes, Department of Psychiatry, School of Medicine, University of Oviedo, CIBERSAM, Oviedo, Spain; Dr Vladimir Carli, National Centre for Suicide Research and Prevention of Mental Ill-Health (NASP), Karolinska Institute, Stockholm, Sweden; Professor Gil Zalsman, Department of Child and Adolescent Psychiatry at Geha Hospital, Tel Aviv University, Tel Aviv, Israel; Professor Philippe Courtet, Head of the Department of Emergency Psychiatry at the Academic Hospital, Montpellier, France; Dr Giuseppe Fanelli, Department of Biomedical and NeuroMotor Sciences, University of Bologna, Bologna, Italy; Professor Zoltan Rihmer, Faculty of Medicine, Department of Clinical and Theoretical Mental Health and Department of Psychiatry and Psychotherapy, Semmelweis University, Budapest, Hungary; Professor Dan Rujescu, Molecular and Clinical Neurobiology, Department of Psychiatry, University of Munich, Munich, Germany; Professor Pilar A. Saiz, Department of Psychiatry, School of Medicine, University of Oviedo, CIBERSAM, Oviedo, Spain; Professor Marco Sarchiapone, Department of Health Sciences, University of Molise, Campobasso, Italy; Professor Alessandro Serretti, Department of Biomedical and NeuroMotor Sciences, University of Bologna, Bologna, Italy; Professor Danuta Wasserman, National Centre for Suicide Research and Prevention of Mental Ill-Health (NASP), Karolinska Institute, Stockholm, Sweden; Professor Jerzy Wasserman, National Centre for Suicide Research and Prevention of Mental Ill-Health (NASP), Karolinska Institute, Stockholm, Sweden; Professor Avi Weizman, Felsenstein Medical Research Center (FMRC), Tel Aviv University, Petah Tikva, Israel.

We thank the US National Institute of Mental Health (NIMH) for providing access to data on the STAR-D, STEP-BD, and CATIE samples. We also thank the authors of previous publications in these datasets, and foremost, we thank the patients and their families who agreed to be enrolled in the studies. Data were obtained from the limited access datasets distributed from the NIH-supported STAR*D (NIMH Contract No. N01MH90003, ClinicalTrials.gov identifier is NCT00021528), STEP-BD (NIMH Contract No. N01MH8001), and CATIE (NIMH Contract No. N01MH900001).

We thank the European Group for the Study of Resistant Depression (GRSD) for providing access to the dataset they collected, as well as the researchers of the consortia that provided the GWAS summary statistics used in our analyses and the participants of the cohorts to which they refer.

## References

Aoki C, Venkatesan C, Go CG, Forman R, Kurose H. 1998. Cellular and subcellular sites for noradrenergic action in the monkey dorsolateral prefrontal cortex as revealed by the immunocytochemical localization of noradrenergic receptors and axons. Cereb Cortex 8(3):269–277.

Arsenault-Lapierre G, Kim C, Turecki G. 2004. Psychiatric diagnoses in 3275 suicides: a meta-analysis. BMC Psychiatry 4:37.

Artero S, Astruc B, Courtet P, Ritchie K. 2006. Life-time history of suicide attempts and coronary artery disease in a community-dwelling elderly population. Int J Geriatr Psychiatry 21(2):108–112.

Bulik-Sullivan B, Finucane HK, Anttila V, Gusev A, Day FR, Loh PR, ReproGen C, Psychiatric Genomics C, Genetic Consortium for Anorexia Nervosa of the Wellcome Trust Case Control C, Duncan L, Perry JR, Patterson N, Robinson EB, Daly MJ, Price AL, Neale BM. 2015. An atlas of genetic correlations across human diseases and traits. Nat Genet 47(11):1236–1241.

Calati R, Ferrari C, Brittner M, Oasi O, Olie E, Carvalho AF, Courtet P. 2019. Suicidal thoughts and behaviors and social isolation: A narrative review of the literature. J Affect Disord 245:653–667.

Choi SW, O’Reilly PF. 2019. PRSice-2: Polygenic Risk Score software for biobank-scale data. Gigascience 8(7).

Cristancho P, O’Connor B, Lenze EJ, Blumberger DM, Reynolds CF, 3rd, Dixon D, Mulsant BH. 2017. Treatment Emergent Suicidal Ideation in depressed older adults. Int J Geriatr Psychiatry 32(6):596–604.

Dennehy EB, Marangell LB, Allen MH, Chessick C, Wisniewski SR, Thase ME. 2011. Suicide and suicide attempts in the Systematic Treatment Enhancement Program for Bipolar Disorder (STEP-BD). J Affect Disord 133(3):423–427.

Dold M, Bartova L, Mendlewicz J, Souery D, Serretti A, Porcelli S, Zohar J, Montgomery S, Kasper S. 2018. Clinical correlates of augmentation/combination treatment strategies in major depressive disorder. Acta Psychiatr Scand 137(5):401–412.

Fanelli G, Benedetti F, Kasper S, Zohar J, Souery D, Montgomery S, Albani D, Forloni G, Ferentinos P, Rujescu D, Mendlewicz J, Serretti A, Fabbri C. 2020. Higher polygenic risk scores for schizophrenia may be suggestive of treatment non-response in major depressive disorder. Prog Neuropsychopharmacol Biol Psychiatry:110170.

Fanelli G, Serretti A. 2019. The influence of the serotonin transporter gene 5-HTTLPR polymorphism on suicidal behaviors: a meta-analysis. Prog Neuropsychopharmacol Biol Psychiatry 88:375–387.

Finucane HK, Bulik-Sullivan B, Gusev A, Trynka G, Reshef Y, Loh PR, Anttila V, Xu H, Zang C, Farh K, Ripke S, Day FR, ReproGen C, Schizophrenia Working Group of the Psychiatric Genomics C, Consortium R, Purcell S, Stahl E, Lindstrom S, Perry JR, Okada Y, Raychaudhuri S, Daly MJ, Patterson N, Neale BM, Price AL. 2015. Partitioning heritability by functional annotation using genome-wide association summary statistics. Nat Genet 47(11):1228–1235.

Friedman RA. 2014. Antidepressants’ black-box warning--10 years later. N Engl J Med 371(18):1666–1668.

Fu Q, Heath AC, Bucholz KK, Nelson EC, Glowinski AL, Goldberg J, Lyons MJ, Tsuang MT, Jacob T, True MR, Eisen SA. 2002. A twin study of genetic and environmental influences on suicidality in men. Psychol Med 32(1):11–24.

Garcia-Gonzalez J, Tansey KE, Hauser J, Henigsberg N, Maier W, Mors O, Placentino A, Rietschel M, Souery D, Zagar T, Czerski PM, Jerman B, Buttenschon HN, Schulze TG, Zobel A, Farmer A, Aitchison KJ, Craig I, McGuffin P, Giupponi M, Perroud N, Bondolfi G, Evans D, O’Donovan M, Peters TJ, Wendland JR, Lewis G, Kapur S, Perlis R, Arolt V, Domschke K, Major Depressive Disorder Working Group of the Psychiatric Genomic C, Breen G, Curtis C, Sang-Hyuk L, Kan C, Newhouse S, Patel H, Baune BT, Uher R, Lewis CM, Fabbri C. 2017. Pharmacogenetics of antidepressant response: A polygenic approach. Prog Neuropsychopharmacol Biol Psychiatry 75:128–134.

Gijzen MWM, Rasing SPA, Creemers DHM, Smit F, Engels R, De Beurs D. 2021. Suicide ideation as a symptom of adolescent depression. a network analysis. J Affect Disord 278:68–77.

Higgins JP, Thompson SG, Deeks JJ, Altman DG. 2003. Measuring inconsistency in meta-analyses. BMJ 327(7414):557–560.

Howland RH. 2008. Sequenced Treatment Alternatives to Relieve Depression (STAR*D). Part 1: study design. J Psychosoc Nurs Ment Health Serv 46(9):21–24.

Laje G, Paddock S, Manji H, Rush AJ, Wilson AF, Charney D, McMahon FJ. 2007. Genetic markers of suicidal ideation emerging during citalopram treatment of major depression. Am J Psychiatry 164(10):1530–1538.

Li Y, Willer CJ, Ding J, Scheet P, Abecasis GR. 2010. MaCH: using sequence and genotype data to estimate haplotypes and unobserved genotypes. Genet Epidemiol 34(8):816–834.

Lu Q, Li B, Ou D, Erlendsdottir M, Powles RL, Jiang T, Hu Y, Chang D, Jin C, Dai W. 2017. A powerful approach to estimating annotation-stratified genetic covariance via GWAS summary statistics. The American Journal of Human Genetics 101(6):939–964.

Luoma JB, Martin CE, Pearson JL. 2002. Contact with mental health and primary care providers before suicide: a review of the evidence. Am J Psychiatry 159(6):909–916.

Marangell LB, Dennehy EB, Wisniewski SR, Bauer MS, Miyahara S, Allen MH, Martinez M, Al Jurdi RK, Thase ME. 2008. Case-control analyses of the impact of pharmacotherapy on prospectively observed suicide attempts and completed suicides in bipolar disorder: findings from STEP-BD. J Clin Psychiatry 69(6):916–922.

Moazzami K, Dolmatova EV, Feurdean M. 2018. Suicidal ideation among adults with cardiovascular disease: The National Health and Nutrition Examination Survey. Gen Hosp Psychiatry 51:5–9.

Mullins N, Bigdeli TB, Borglum AD, Coleman JRI, Demontis D, Mehta D, Power RA, Ripke S, Stahl EA, Starnawska A, Anjorin A, M.R.C. Psych, Corvin A, Sanders AR, Forstner AJ, Reif A, Koller AC, Swiatkowska B, Baune BT, Muller-Myhsok B, Penninx B, Pato C, Zai C, Rujescu D, Hougaard DM, Quested D, Levinson DF, Binder EB, Byrne EM, Agerbo E, Dr.Med.Sc, Streit F, Mayoral F, Bellivier F, Degenhardt F, Breen G, Morken G, Turecki G, Rouleau GA, Grabe HJ, Volzke H, Jones I, Giegling I, Agartz I, Melle I, Lawrence J, M.R.C. Psych, Walters JTR, Strohmaier J, Shi J, Hauser J, Biernacka JM, Vincent JB, Kelsoe J, Strauss JS, Lissowska J, Pimm J, M.R.C. Psych, Smoller JW, Guzman-Parra J, Berger K, Scott LJ, Jones LA, Azevedo MH, Trzaskowski M, Kogevinas M, Rietschel M, Boks M, Ising M, Grigoroiu-Serbanescu M, Hamshere ML, Leboyer M, Frye M, Nothen MM, Alda M, Preisig M, Nordentoft M, Boehnke M, O’Donovan MC, Owen MJ, Pato MT, Renteria ME, Budde M, Dipl P, Weissman MM, Wray NR, Bass N, M.R.C. Psych, Craddock N, Smeland OB, Andreassen OA, Mors O, Gejman PV, Sklar P, McGrath P, Hoffmann P, McGuffin P, Lee PH, Mortensen PB, Kahn RS, Ophoff RA, Adolfsson R, Van der Auwera S, Djurovic S, Kloiber S, Heilmann-Heimbach S, Jamain S, Hamilton SP, McElroy SL, Lucae S, Cichon S, Schulze TG, Hansen T, Werge T, Air TM, Nimgaonkar V, Appadurai V, Cahn W, Milaneschi Y, Major Depressive Disorder Working Group of the Psychiatric Genomics C, Bipolar Disorder Working Group of the Psychiatric Genomics C, Schizophrenia Working Group of the Psychiatric Genomics C, Fanous AH, Kendler KS, McQuillin A, Lewis CM. 2019. GWAS of Suicide Attempt in Psychiatric Disorders and Association With Major Depression Polygenic Risk Scores. Am J Psychiatry 176(8):651–660.

Mullins N, Perroud N, Uher R, Butler AW, Cohen-Woods S, Rivera M, Malki K, Euesden J, Power RA, Tansey KE, Jones L, Jones I, Craddock N, Owen MJ, Korszun A, Gill M, Mors O, Preisig M, Maier W, Rietschel M, Rice JP, Muller-Myhsok B, Binder EB, Lucae S, Ising M, Craig IW, Farmer AE, McGuffin P, Breen G, Lewis CM. 2014. Genetic relationships between suicide attempts, suicidal ideation and major psychiatric disorders: a genome-wide association and polygenic scoring study. Am J Med Genet B Neuropsychiatr Genet 165B(5):428–437.

Nock MK, Hwang I, Sampson N, Kessler RC, Angermeyer M, Beautrais A, Borges G, Bromet E, Bruffaerts R, de Girolamo G, de Graaf R, Florescu S, Gureje O, Haro JM, Hu C, Huang Y, Karam EG, Kawakami N, Kovess V, Levinson D, Posada-Villa J, Sagar R, Tomov T, Viana MC, Williams DR. 2009. Cross-national analysis of the associations among mental disorders and suicidal behavior: findings from the WHO World Mental Health Surveys. PLoS Med 6(8):e1000123.

Palla L, Dudbridge F. 2015. A Fast Method that Uses Polygenic Scores to Estimate the Variance Explained by Genome-wide Marker Panels and the Proportion of Variants Affecting a Trait. Am J Hum Genet 97(2):250–259.

Perlis RH, Purcell S, Fava M, Fagerness J, Rush AJ, Trivedi MH, Smoller JW. 2007. Association between treatment-emergent suicidal ideation with citalopram and polymorphisms near cyclic adenosine monophosphate response element binding protein in the STAR*D study. Arch Gen Psychiatry 64(6):689–697.

Rivero G, Gabilondo AM, Garcia-Sevilla JA, La Harpe R, Callado LF, Meana JJ. 2014. Increased alpha2-and beta1-adrenoceptor densities in postmortem brain of subjects with depression: differential effect of antidepressant treatment. J Affect Disord 167:343–350.

Ruderfer DM, Walsh CG, Aguirre MW, Tanigawa Y, Ribeiro JD, Franklin JC, Rivas MA. 2020. Significant shared heritability underlies suicide attempt and clinically predicted probability of attempting suicide. Mol Psychiatry 25(10):2422–2430.

Sachs GS, Thase ME, Otto MW, Bauer M, Miklowitz D, Wisniewski SR, Lavori P, Lebowitz B, Rudorfer M, Frank E, Nierenberg AA, Fava M, Bowden C, Ketter T, Marangell L, Calabrese J, Kupfer D, Rosenbaum JF. 2003. Rationale, design, and methods of the systematic treatment enhancement program for bipolar disorder (STEP-BD). Biol Psychiatry 53(11):1028–1042.

Solmi M, Veronese N, Galvano D, Favaro A, Ostinelli EG, Noventa V, Favaretto E, Tudor F, Finessi M, Shin JI, Smith L, Koyanagi A, Cester A, Bolzetta F, Cotroneo A, Maggi S, Demurtas J, De Leo D, Trabucchi M. 2020. Factors Associated With Loneliness: An Umbrella Review Of Observational Studies. J Affect Disord 271:131–138.

Stahl SM, Stahl SM. 2013. Stahl’s essential psychopharmacology: neuroscientific basis and practical applications. Cambridge university press.

Stickley A, Koyanagi A, Ueda M, Inoue Y, Waldman K, Oh H. 2020. Physical multimorbidity and suicidal behavior in the general population in the United States. J Affect Disord 260:604–609.

Strang F, Schunkert H. 2014. C-reactive protein and coronary heart disease: all said--is not it? Mediators Inflamm 2014:757123.

Stroup TS, McEvoy JP, Swartz MS, Byerly MJ, Glick ID, Canive JM, McGee MF, Simpson GM, Stevens MC, Lieberman JA. 2003. The National Institute of Mental Health Clinical Antipsychotic Trials of Intervention Effectiveness (CATIE) project: schizophrenia trial design and protocol development. Schizophr Bull 29(1):15–31.

Turecki G, Brent DA, Gunnell D, O’Connor RC, Oquendo MA, Pirkis J, Stanley BH. 2019. Suicide and suicide risk. Nat Rev Dis Primers 5(1):74.

Voracek M, Loibl LM. 2007. Genetics of suicide: a systematic review of twin studies. Wien Klin Wochenschr 119(15-16):463–475.

WHO WHO. 2019. World health statistics 2019: monitoring health for the SDGs, sustainable development goals.

Yanowitch R, Coccaro EF. 2011. The neurochemistry of human aggression. Adv Genet 75:151–169.

Zheutlin AB, Dennis J, Karlsson Linner R, Moscati A, Restrepo N, Straub P, Ruderfer D, Castro VM, Chen CY, Ge T, Huckins LM, Charney A, Kirchner HL, Stahl EA, Chabris CF, Davis LK, Smoller JW. 2019. Penetrance and Pleiotropy of Polygenic Risk Scores for Schizophrenia in 106,160 Patients Across Four Health Care Systems. Am J Psychiatry 176(10):846–855.

Zhong Y, Pham S, Porta G, Douaihy A, Marsland A, Brent D, Melhem NM. 2020. Increased burden of cardiovascular risk among youth suicide attempters. Psychol Med:1–9.

