## Supplementary Figure 1 for "Polygenic risk scores for psychiatric, inflammatory, and cardio-metabolic traits and diseases highlight possible genetic overlaps with suicide attempt and treatment-emergent suicidal ideation"

**Fig. S1.** Forest plot showing the association between the polygenic risk score for major depressive disorder (calculated at the genome-wide P-threshold of 0.05) and **suicide** **attempt** across the four included clinical cohorts. Abbreviations: FE, fixed effects; CI, confidence intervals; SE, standard error; CATIE, Clinical Antipsychotic Trials of Intervention Effectiveness; GSRD, European Group for the Study of Resistant Depression; STAR*D, Sequenced Treatment Alternatives to Relieve Depression, STEP-BD, Systematic Treatment Enhancement Program for Bipolar Disorder.

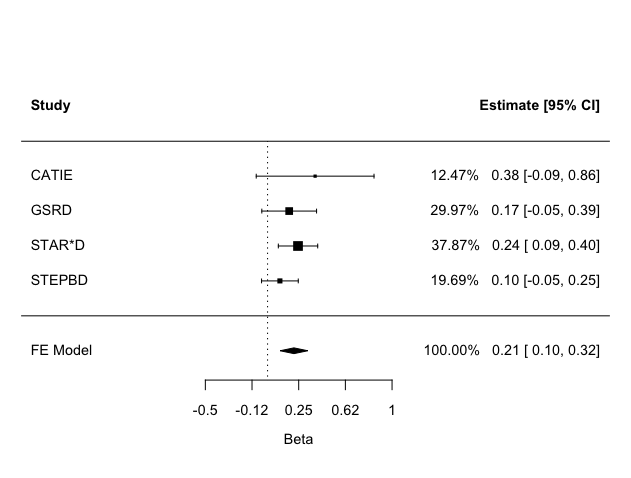

Test for overall effect: **Beta = 0.212, SE = 0.057, Z = 3.749, P = 1.77e-4**
